# Baseline Characteristics of Black and Hispanic Adults with Hypertension in Healthy Food Priority Areas: The THRIVE Food-is-Medicine Pilot Trial

**DOI:** 10.1101/2025.09.22.25336404

**Authors:** India Washington, Mojisola Olusola-Bello, D’Janee Kyeremeh, Irma Iribe, Adeline Assani-Uva, Janice Dugbartey, Oluwatosin Tomiwa, Maricielo Leasure, Elohor Oborevwori, Aminata Sinyan, Adrián McMahon, Maya Kramer-Johansen, Peiyu Chen, Christy Rodriguez, William Xiao, Samuel Gledhill, Samuel Yeboah-Manson, Lydia Vassiliadi, Shanshan Song, Meng Yuan, Chelsea Akubo, Cassie Romero, Jennifer Freeman, Anna Maria Izquierdo-Porrera, Valerie K. Sullivan, Yvonne Commodore-Mensah, Oluwabunmi Ogungbe, the THRIVE Food-is-Medicine Team

## Abstract

**Background:** Hypertension disproportionately affects Black and Hispanic adults, with disparities amplified by structural barriers, including limited access to healthy food. Understanding cardiovascular risk profiles is essential for designing effective interventions. We assessed baseline characteristics of participants in the THRIVE Food-is-Medicine pilot trial.

**Methods:** This baseline analysis included 80 Black and Hispanic adults from Maryland participating in a randomized controlled trial of a culturally tailored Food-is-Medicine intervention. Participants had hypertension or uncontrolled blood pressure (≥120/80 mmHg), and resided in census tracts with low Healthy Food Availability Index scores. We collected sociodemographic characteristics, clinical measures, laboratory values, and dietary patterns. Descriptive statistics were calculated using means (±standard deviation) for continuous variables and frequencies (percentages) for categorical variables.

**Results:** Participants had high educational attainment (45% ≥ master’s degree), yet economic vulnerabilities persisted: 70% <$50,000 annually, 28% unemployed, and 31% lacked health insurance. Food insecurity affected 36%. Mean systolic and diastolic blood pressure were 138.1±16.1 and 82.6±10.3 mmHg, respectively. While 45% had Stage 2 hypertension, only 58% owned blood pressure monitors. Regarding dietary quality, the mean DASH adherence score was 3.83±1.9 out of 9 points, with 58% below 4.5. Potassium intake was low; only 29% met DASH potassium targets.

**Conclusion:** Based on these data, THRIVE participants faced intersecting vulnerabilities, including economic insecurity, limited healthcare access, food insecurity, and poor dietary quality. This profile shows the multilayered drivers of hypertension disparities and substantial cardiovascular risk burden. Findings will guide intervention refinement of larger-scale nutrition and food-is-medicine trials addressing hypertension disparities in under-resourced communities.

## INTRODUCTION

Hypertension affects nearly half of U.S. adults and remains the leading modifiable risk factor for cardiovascular disease, stroke, and premature mortality.^1,2^ Despite significant advances in treatment, racial and ethnic disparities persist: Black and Hispanic adults have higher hypertension rates and poorer blood pressure control than White adults, contributing to ongoing cardiovascular health inequities.^3–5^

The cardiovascular burden among racial and ethnic minorities is amplified by multilayered interactions between social drivers of health, including economic insecurity, limited healthcare access, and structural barriers to healthy lifestyle adoption.^6,7^ Among these drivers, food insecurity and limited access to nutritious foods are fundamental and are modifiable factors influencing cardiovascular health.^8,9^ The Dietary Approaches to Stop Hypertension (DASH) dietary pattern, which emphasizes fruits, vegetables, whole grains, and lean proteins while limiting sodium and processed foods, has shown significant blood pressure-lowering effects.^10–12^ However, adherence to DASH remains suboptimal, particularly among Black and Hispanic populations who face greater barriers to accessing and affording DASH-aligned foods.^13,14^

Healthy Food Priority Areas (HFPAs), previously termed “food deserts,” are characterized by limited availability of affordable, nutritious foods and disproportionately affect low-income communities and communities of color.^15,16^ Residents of HFPAs face barriers, including inadequate access to grocery stores, higher costs for healthy foods, limited transportation, and reduced availability of culturally appropriate produce, all factors that can hinder the adoption of heart-healthy dietary patterns.^15,17^

Recognizing these interconnected challenges, Food-is-Medicine interventions have emerged as promising approaches to address both food access barriers and cardiovascular health disparities simultaneously.^18^ Food-is-Medicine programs, which often include produce prescriptions, medically tailored groceries, medically tailored meals, and nutrition education, aim to improve health outcomes by addressing food insecurity and promoting access to nutritious foods.^19^ Successful implementation of Food-is-Medicine interventions requires an understanding of the cardiovascular risk profiles and social determinants affecting target populations.^20^ This is important for tailoring interventions to address specific community needs and identifying high-risk groups. Moreover, characterizing the intersection between socioeconomic factors, food access, dietary quality, and cardiovascular risk can inform broader policy initiatives aimed at addressing structural determinants of health disparities.^8,21^

The THRIVE (AdapTive personalized dietitian coacHing, messaging, and pRoduce prescrIption to improVE healthy dietary behaviors) pilot trial is a Food-is-Medicine intervention designed specifically for Black and Hispanic adults with hypertension residing in Maryland HFPAs.^22^ Developed using human-centered design principles in partnership with community stakeholders, clinicians, and target populations, THRIVE centers cultural relevance and community acceptability.^23^ The THRIVE intervention combines produce prescriptions, culturally tailored nutrition education, dietitian coaching, and linkages to social resources, addressing multiple barriers to hypertension management simultaneously. The objective of this study was to assess the baseline sociodemographic characteristics, cardiovascular risk factors, dietary patterns, and social determinants of health among Black and Hispanic adults with hypertension enrolled in the THRIVE pilot study. These findings will contribute to the growing evidence base on cardiovascular health disparities and inform strategies for implementing effective Food-is-Medicine interventions in underserved communities with high cardiovascular risk burden.

### Registration

URL: https://www.clinicaltrials.gov; Unique identifier: NCT06257550.

## NOVELTY AND RELEVANCE

### What Is New?

This is the first study to describe baseline sociodemographic, clinical, and dietary characteristics of Black and Hispanic adults with hypertension living in Healthy Food Priority Areas and enrolled in a culturally tailored Food-is-Medicine intervention. We document poor DASH diet adherence, high rates of food and nutrition insecurity, and gaps in blood pressure self-monitoring, despite high educational attainment.

### What Is Relevant?

Our findings highlight the multilayered drivers of hypertension disparities, including economic insecurity, limited healthcare access, and structural barriers to healthy eating. These data provide the foundation for designing and scaling Food-is-Medicine interventions that are culturally tailored and community-centered.

### Clinical/Pathophysiological Implications?

By characterizing cardiovascular risk factors and social determinants in a high-risk, under-resourced population, this study underscores the need to integrate nutrition security interventions into hypertension management and policy.

These findings can inform clinical practice and health system efforts to reduce disparities in blood pressure control and cardiovascular outcomes.

## METHODS

### Study Design

This baseline analysis examines data from participants enrolled in the THRIVE pilot trial,^22^ a two-arm randomized pilot feasibility trial testing a Food-is-Medicine produce prescription intervention among Black and Hispanic adults with hypertension. The study was conducted in Montgomery and Prince George’s Counties, Maryland, within census tracts designated as Healthy Food Priority Areas (HFPAs) by the Montgomery County Department of Planning.^24^

### Study Population

Participants were required to be 18 years or older and self-identify as Black/African American or Hispanic. Blood pressure measurements had to fall within the range of 120-139/80-89 mm Hg or exceed 140/90 mm Hg, as determined by a validated Omron blood pressure device. Additionally, eligible individuals had to reside in census tracts designated by the Montgomery County Department of Planning as HFPA, characterized by a low Healthy Food Availability Index score (0-9.5), a median household income at or below 185% of the Federal Poverty Level, and over 30% of households lacking vehicle access with a supermarket distance exceeding a quarter mile.^17^ Participants were also required to have refrigeration and essential food appliances such as a microwave or stove, along with a cell phone to receive messages.

Individuals under 18 years of age, those diagnosed with Type 1 or Type 2 diabetes (hemoglobin A1c ≥6.5% or receiving diabetes treatment), and those with end-stage renal disease (ESRD) were excluded from this study. Other exclusion factors included conditions that could interfere with outcome measurement, such as dialysis, or the presence of serious medical conditions like cancer that limit life expectancy or require active management. Furthermore, participants with significant food allergies, dietary restrictions affecting adherence, cognitive impairments preventing participation, or those currently engaged in a care management program related to health conditions such as weight reduction or smoking cessation were excluded. Additionally, individuals already enrolled in another clinical trial that might interfere with the THRIVE study, those planning to relocate outside the geographic area within 12 months, and those unwilling to provide informed consent were deemed ineligible.

### Patient Recruitment and Randomization

Participants were recruited from May to December 2024 from eight different community sites within HFPAs in Maryland. Recruitment methods included: (1) outreach through clinic partners, i.e. Care for Your Health clinic^25^ sites and mobile “pop-up” clinics in pre-identified neighborhoods; (2) partnerships with faith-based organizations^26,27^ in Black and Hispanic communities; (3) targeted social media advertising on Facebook and Instagram within specific zip codes; (4) community flyers and website-based sign-up; and (5) referrals from primary care physicians and community partners. Over 1,000 people were approached at the sites, and 230 signed up for the study.

Interested individuals completed pre-screening via online form on StudyPages^28^ or a telephone interview. Eligible participants attended an in-person screening visit at our clinic and community locations for final eligibility confirmation, informed consent, and baseline data collection. The screening visit included blood pressure measurement, detailed eligibility screening, and written informed consent in English, Spanish, Creole, French, or Twi. The study protocol was approved by the Johns Hopkins University Institutional Review Board (IRB00427492) and registered at ClinicalTrials.gov (NCT06257550).

### Human-Centered Design Process

Before participant recruitment, the THRIVE intervention was co-designed through a human-centered design process involving 36 community stakeholders (29 female, 6 male, 17 English-speaking, 18 Spanish-speaking/bilingual) in three iterative design sessions.^23^ Participants included community residents with hypertension, healthcare providers, local food system representatives, and community organization leaders. Using social cognitive theory and the human-centered design double-diamond framework, the sessions followed this approach: (1) orientation and listening (virtual session), (2) prototyping and feedback (in-person), and (3) process mapping and refinement (in-person). Key themes that emerged included the need for personalized dietitian support with cultural competency, flexible produce prescription programs with practical nutrition education, and peer support networks. This co-design process informed intervention components, delivery procedures, and implementation strategies to ensure cultural relevance and community acceptability.

### THRIVE Pilot Study Intervention

Following the co-design sessions, the THRIVE intervention was refined to meet community needs. The intervention includes multiple culturally tailored components designed to reduce barriers to healthy eating and improve adherence to the DASH dietary pattern in populations residing within HFPAs. Key elements of the intervention include personalized dietitian coaching, guiding grocery habits and DASH goals, weekly culturally-tailored adaptive messaging, produce prescription via FARMacy Market, linkages to social services based on identified needs, adherence support strategies, and interventionist training with an emphasis on culturally relevant recipes and food traditions.

The intervention was designed to address access, education, personalization, and cultural barriers to healthy eating. Participants received weekly produce prescriptions valued at $35, fulfilled through Community FarmShare’s^29^ “FARMacy” mobile market at strategic locations. The market was stocked with seasonal, culturally appropriate produce from 12+ local farms, with selections aligned to DASH dietary principles and participants’ food preferences identified during dietitian coaching sessions. Registered dietitians with expertise in hypertension management provided individualized nutrition counseling through an initial assessment (30-45 minutes) covering dietary patterns, cultural food traditions, and collaborative goal-setting, followed by bi-weekly sessions (15-20 minutes) for progress monitoring, barrier identification, and recipe adaptation. Communication was delivered flexibly through telehealth, secure messaging, and phone consultations, and emphasized the incorporation of traditional foods into DASH-compliant meal plans. An intelligent messaging system delivered personalized nutrition education through weekly pulse surveys assessing produce utilization and dietary adherence, triggering automated, culturally-relevant nutrition tips and educational content, including infographics and recipe cards featuring traditional dishes modified for DASH compliance. The intervention also included social needs assessment using validated screening tools, with referrals to community resources facilitated through The MEDI platform,^30^ food safety education, and flexible scheduling to accommodate participant barriers.

The comparator group received standard produce bags from Community FarmShare without additional coaching.

### Intervention Fidelity

To study registered dietitians who had received training in DASH principles, motivational interviewing, and cultural competency before participant contact. Fidelity was monitored through documentation of all dietitian-participant interactions in StudyPages using standardized forms. To prevent treatment contamination, control group participants received standard produce bags through home delivery and were not exposed to the FARMacy. All study staff were trained to maintain group allocation confidentiality and avoid discussing intervention-specific content with control participants. Intervention adherence was tracked through dietitian session attendance rates, produce prescription utilization via FARMacy market records, and weekly pulse survey response rates.

### Study Measures

#### Sociodemographic and Social Needs Factors

Baseline sociodemographic data were collected via standardized questionnaires, including age, sex, race/ethnicity, education level, employment status, household income, insurance status, country of birth, years in the United States, and housing stability. Social needs were assessed using the Accountable Health Communities Health-Related Social Needs Screening Tool.^31^ Food security was evaluated using the USDA Six-Item Short Form Food Security Survey Module.^32^ Nutrition security was evaluated using the GusNIP Nutrition Security Screener.^33^ Healthcare utilization patterns were assessed using instruments from the PhenX toolkit.^34^

##### Clinical Assessments

Trained research staff conducted standardized clinical assessments per protocol. Blood pressure was measured using validated Omron devices after participants rested for ≥5 minutes, with three measurements taken 1 minute apart and the average of the second and third readings recorded. Hypertension staging was per the 2017 American College of Cardiology/American Heart Association guidelines.^35^ Height and weight were measured using calibrated stadiometers and scales, with body mass index calculated as kg/m².

##### Laboratory Measurements

Trained phlebotomists collected fasting blood samples for analysis of hemoglobin A1c, a comprehensive metabolic panel (including glucose, creatinine, electrolytes), and a lipid profile (total cholesterol, LDL cholesterol, HDL cholesterol, and triglycerides). Point-of-care hemoglobin A1c testing was performed using the A1cNow®+ System during screening visits.^36^ Spot urine samples were collected to assess urine sodium and potassium levels. All laboratory analyses were performed by LabCorp and Quest using standard clinical protocols.

##### Dietary Assessment

Dietary intake was assessed using an interviewer-administered 24-hour recall using the Nutrition Data System for Research.^37^ Participants completed multiple 24-hour dietary recalls with assistance from trained research staff as needed. DASH diet adherence was calculated based on nine nutritional targets, per Mellen et al.^38^: total fat, saturated fat, protein, cholesterol, fiber, magnesium, calcium, sodium, and potassium, with scores ranging from 0–9 points. Participants were categorized as having low DASH adherence (score <4.5) or high DASH adherence (score ≥4.5) based on validated cut-points.^39^

##### Additional Assessments

Physical activity was assessed using the Simple Physical Activity Questionnaire.^40^ Sleep quality was measured using the PROMIS Sleep Disturbance Short Form.^41^ Self-efficacy was evaluated using the New General Self-Efficacy Scale.^42^ Health-related quality of life was assessed using the SF-12 Health Survey.^43^ Health literacy was measured using the eHEALS scale.^44^ Smoking and alcohol use were evaluated with the TAPS tool.^45^ Discrimination exposure was measured using the Experiences of Discrimination Instrument,^46^ and depression levels were evaluated with the 8-item Patient Health Questionnaire (PHQ-8).^47^ Social isolation was measured by the De Jong Gierveld Loneliness Scale,^48^ and perceived stress was assessed using the Perceived Stress Scale by Mind Garden.^49^

All questionnaires were administered in English, Spanish, French, Creole, and Twi.

### Randomization

Upon completion of baseline assessments, participants were randomized 1:1 to either the THRIVE intervention or the standard produce bag control group using an automated system with permuted block randomization (block sizes of 4 and 8), stratified by race/ethnicity and baseline blood pressure category. The study team (except 2 designated persons) and outcome assessors were masked to allocation assignment; due to the nature of the intervention, the participants and interventionists (dietitians) could not be masked.

### Statistical Analysis

We conducted the analyses using Stata/SE v18.0 (StataCorp LLC).^50^ We performed descriptive analyses to characterize the baseline profile of THRIVE pilot trial participants and assess randomization balance between treatment groups. Continuous variables were summarized using means and standard deviations, while categorical variables were presented as frequencies and percentages. To evaluate potential differences between the standard produce bag control and THRIVE intervention groups at baseline, we used a two-sample t-test for continuous variables (e.g., age, blood pressure measurements, laboratory values, dietary intake metrics) and chi-square tests for categorical variables (e.g., demographic characteristics, clinical categories, food security status). For variables with small, expected cell counts, Fisher’s exact test was used as appropriate. Statistical significance was defined as two-sided P < 0.05.

DASH diet adherence scores were calculated using validated scoring methods based on nine nutritional targets (total fat, saturated fat, protein, cholesterol, fiber, magnesium, calcium, sodium, and potassium) as outlined by Mellen et al.^38^ Individual nutrient component scores (0-1 points each) were summed to create total DASH scores ranging from 0-9 points, with higher scores indicating greater adherence. Participants were subsequently categorized into adherence groups using these cut-points: low DASH adherence (scores <4.5) versus high DASH adherence (scores ≥4.5).^39^ The distribution of DASH adherence categories between treatment groups was compared using chi-square tests. Individual DASH component scores were calculated for each of the nine nutrients and visualized using radar plots to illustrate adherence patterns across specific dietary targets. We examined the distribution of total DASH scores by treatment group using histograms and assessed the proportion of participants achieving high versus low DASH adherence within each group.

Missing data patterns were assessed and reported for all baseline variables. Variables with substantial missing data (>20%) were excluded from comparative analyses and noted in table footnotes to maintain transparency about data completeness. Notable exclusions included urine creatinine, additional spot urine analytes, and some comprehensive metabolic panel components not directly related to study outcomes due to high missingness. This baseline analysis focused on descriptive characterization and assessment of randomization balance rather than inferential testing of intervention effects, as the primary efficacy analyses will be conducted upon completion of the 24-week follow-up period. All analyses adhered to the intention-to-treat principle based on randomized treatment assignment, ensuring that participants were analyzed according to their original group allocation regardless of adherence or completion status.

## RESULTS

Of 80 enrolled participants, 40 were randomized to the THRIVE intervention and 40 to the standard produce bag control group. The mean age was 54.5±11.4 years.

### Participant Characteristics

Baseline demographic characteristics were generally balanced between groups (**Table 1**). Black/African American participants comprised the largest proportion (66.7% control vs. 57.9% intervention), followed by Hispanic/Mexican American participants (27.8% vs. 39.5%). Small proportions were identified as Asian or Other race across both groups. Income distribution was similar between groups, with approximately two-thirds reporting annual income below $50,000. The control group had slightly higher rates of tobacco use (19.4% vs. 7.9%, P = 0.15) and alcohol use (38.9% vs. 28.9%, P = 0.37), though differences were not statistically significant.

**Table I.**
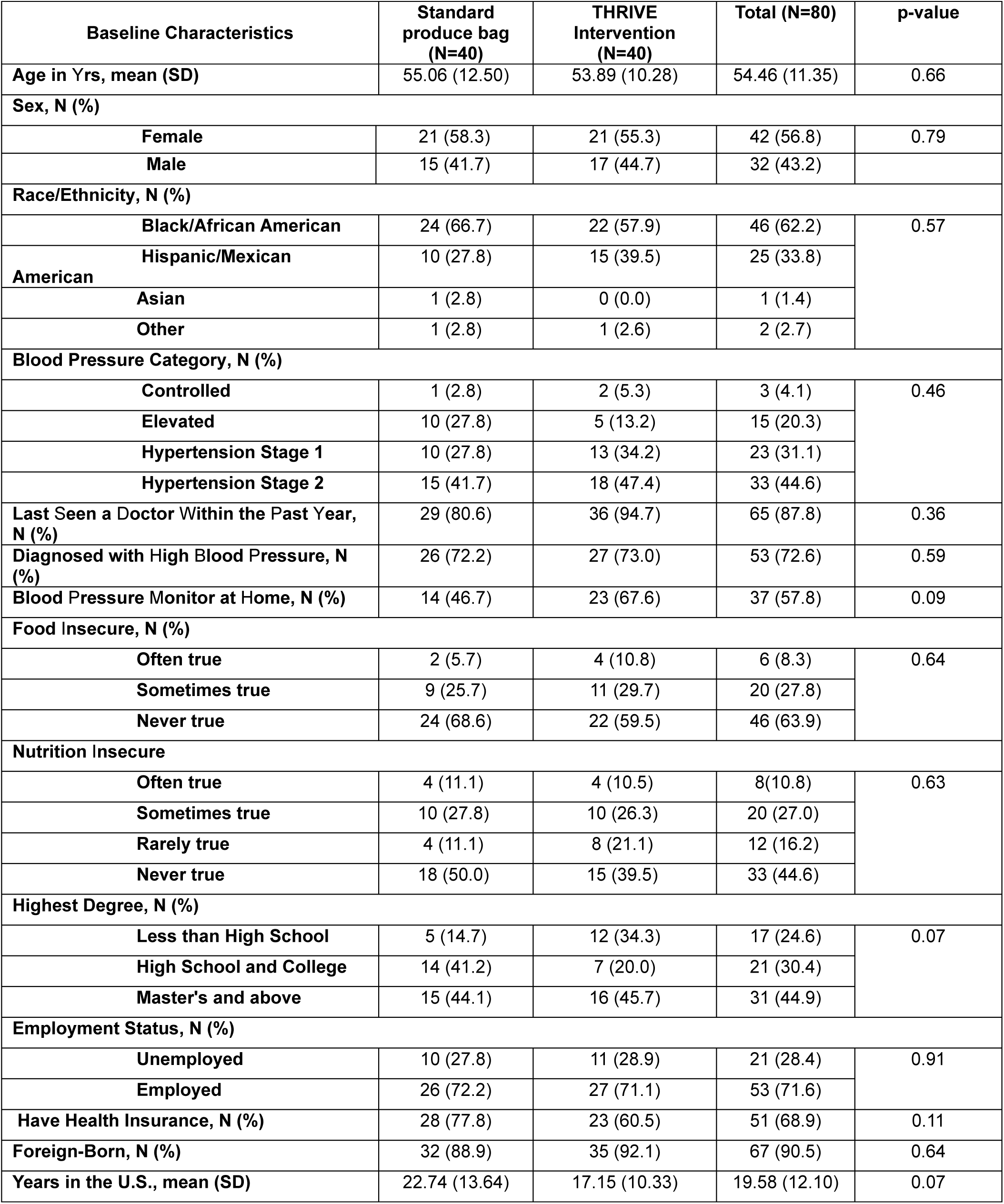
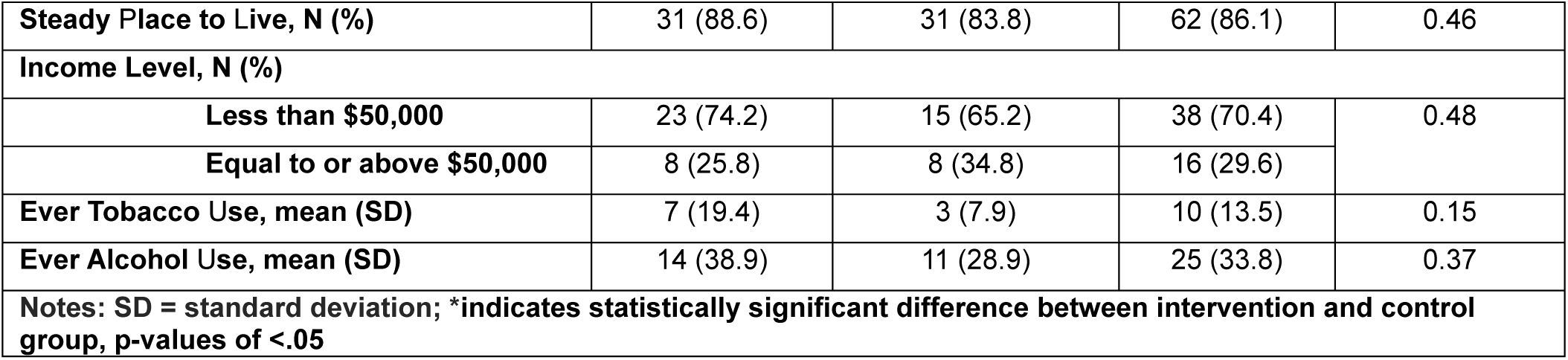
Baseline Sociodemographic Characteristics of THRIVE Pilot Study.

Blood pressure categories did not significantly differ between groups (P = 0.455), with comparable proportions classified as controlled, elevated, hypertension Stage 1, or hypertension Stage 2. Only about 86% reported a steady place to live.

### Healthcare Access and Immigration Background

Most participants had regular healthcare contact, with 88% having seen a doctor within the past year and 73% having a previous diagnosis of hypertension (**Table 1**). However, self-management barriers existed: only 58% of individuals owned home blood pressure monitors, despite the high prevalence of hypertension that requires regular monitoring. Insurance coverage was suboptimal, with 31% of participants lacking health insurance. The sample was predominantly immigrant, with 91% of the participants being foreign-born and an average U.S. residence of 20 years.

### Food and Nutrition Security

Food insecurity affected over one-third of participants (36%), with 8% reporting that food “often did not last” and 28% experiencing this “sometimes” (**Table 1**). Food insecurity rates were similar between groups (P = 0.64), confirming successful randomization on this key intervention target.

### Cardiovascular Risk Factors

Cardiovascular risk factors were generally balanced between groups (**Table 2**). Overall, participants had a substantial cardiovascular risk burden, with a mean systolic/diastolic blood pressure of 138.1±16.1/82.6±10.3 mmHg, and 45% had Stage 2 hypertension. Despite this, only 58% owned home blood pressure monitors, and 73% had previously been diagnosed with hypertension.

**Table II.**
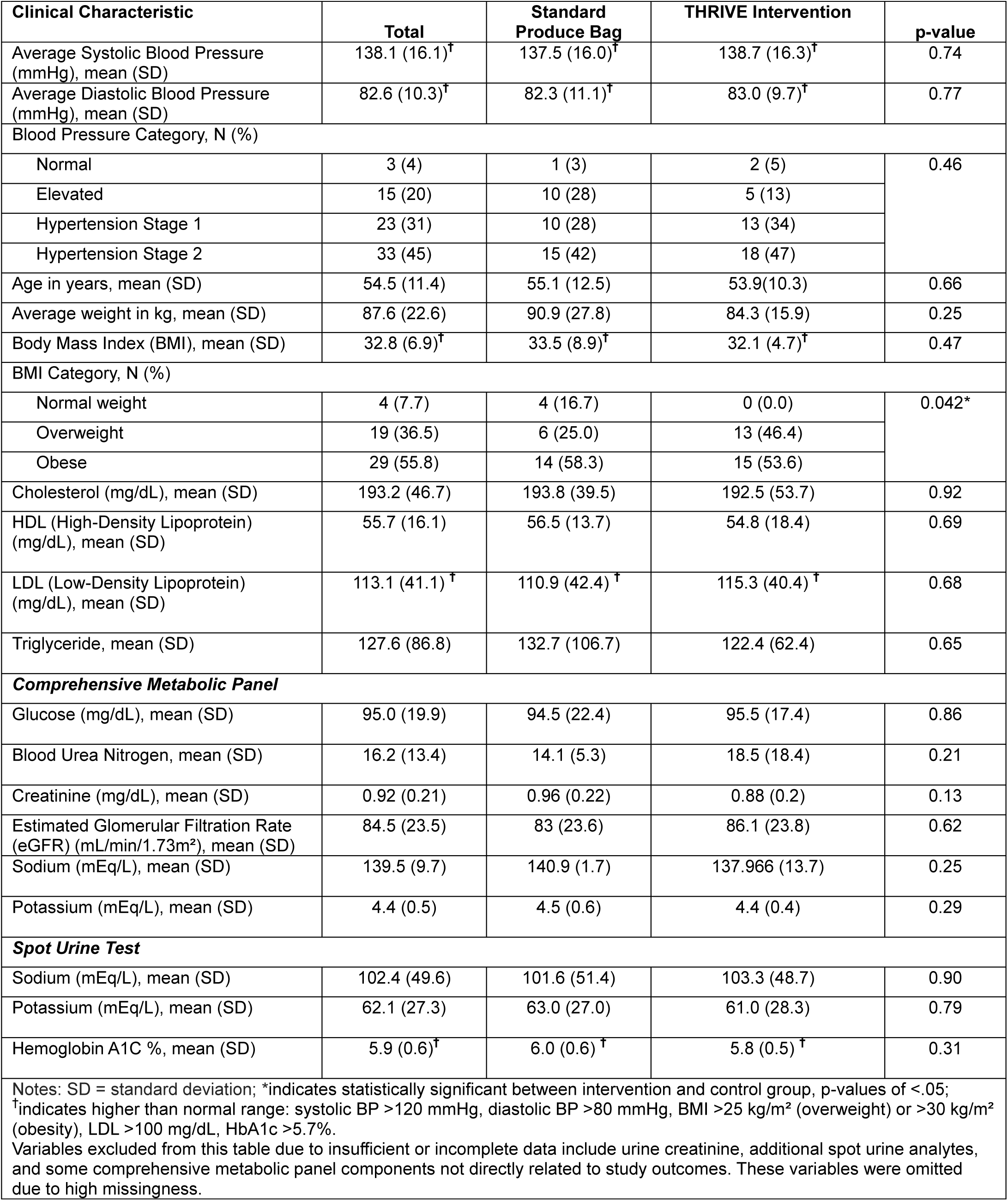
Baseline Clinical Characteristics of THRIVE Pilot Study.

Mean BMI was 32.8±6.9 kg/m², with 56% classified as obese and 37% as overweight. The notable exception in group balance was BMI category distribution (P = 0.042): no THRIVE participants were normal weight compared to 17% of controls, while 46% of THRIVE participants were overweight versus 25% of controls.

Laboratory values showed no significant between-group differences. Mean total cholesterol was 193.2±46.7 mg/dL, HDL 55.7±16.1 mg/dL, LDL 113.1±41.1 mg/dL, and triglycerides 127.6±86.8 mg/dL. Mean HbA1c was 5.9±0.6%. Comprehensive metabolic panel results were within normal ranges: glucose 95.0±19.9 mg/dL, eGFR 84.5±23.5 mL/min/1.73m², creatinine 0.92 ±0.21 mg/dL, serum sodium 139.5±9.7 mEq/L, and potassium 4.4±0.5 mEq/L.

### Dietary Recall Data

Baseline dietary intake patterns were generally comparable between groups, with no significant differences across most nutrient categories (**Table 3**). Both groups had suboptimal dietary quality characterized by high sodium intake (1,839 vs. 1,956 mg, P = 0.67), low potassium intake (1,995 vs. 2,014 mg, P = 0.95), and poor adherence to dietary quality indices. Mean DASH scores were low in both groups (3.7 vs. 3.9 out of 9 possible points, P = 0.74), and Healthy Eating Index scores were moderate (61.3 vs. 60.6 out of 100, P = 0.84).

**Table III.**
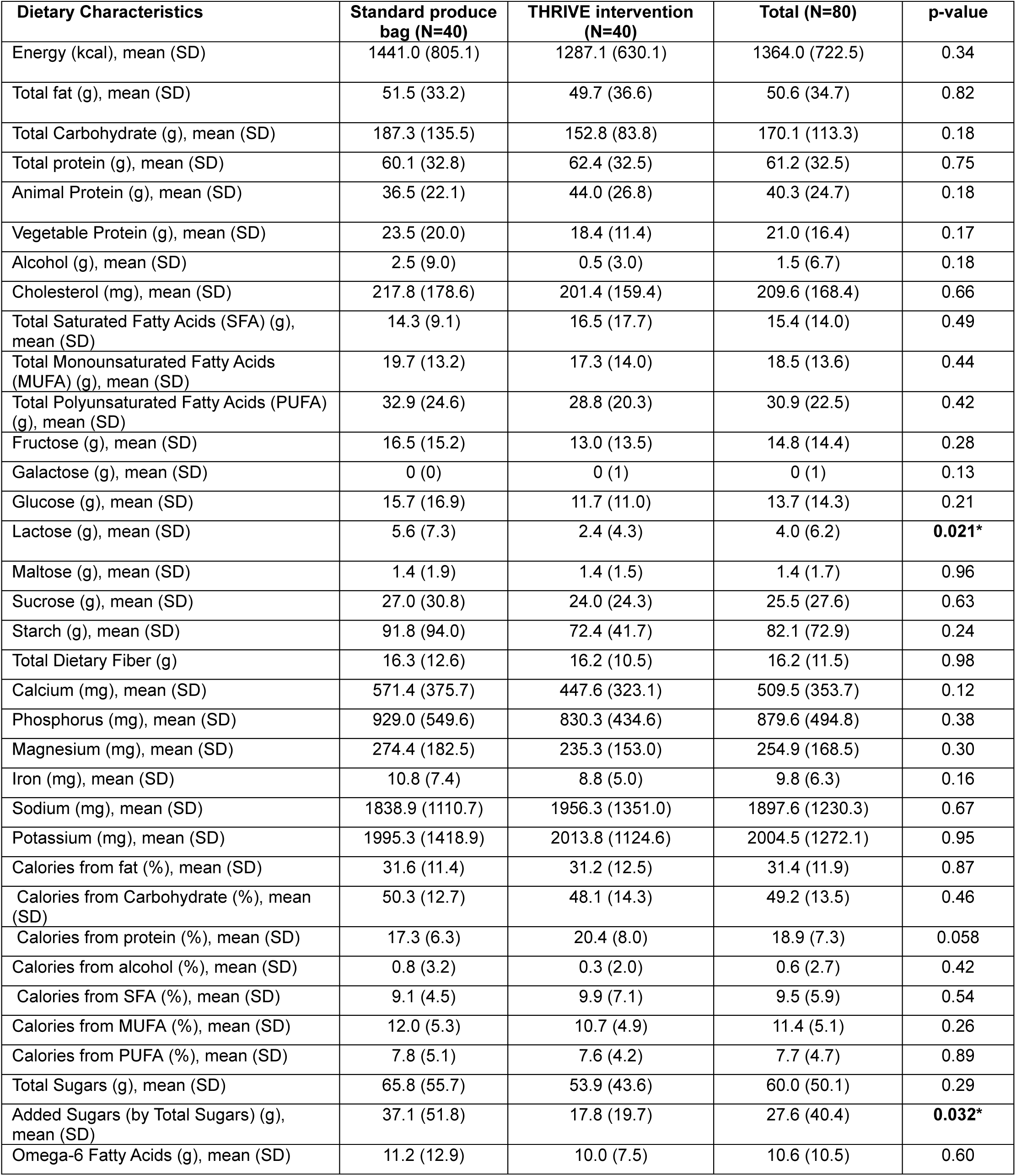
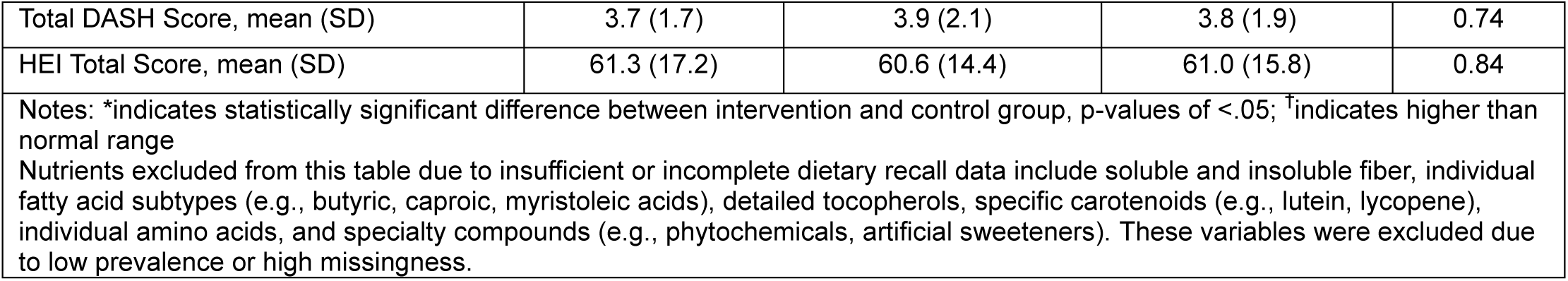
Baseline Dietary Recall Data for THRIVE Pilot Study.

There were two significant between-group differences: the control group had higher lactose intake (5.6 vs. 2.4 g, P = 0.021) and substantially higher added sugar consumption (37.1 vs. 17.8 g, P = 0.032). Mean energy intake was similar between groups (1,441 vs. 1,287 kcal, P = 0.34), with comparable macronutrient distributions— approximately one-third of calories from fat, half from carbohydrates, and the remainder from protein.

Micronutrient intakes were generally adequate but showed no significant between-group differences. The control group reported slightly higher intakes of calcium (571 vs. 448 mg, P = 0.12), magnesium (274 vs. 235 mg, P = 0.30), and iron (10.8 vs. 8.8 mg, P = 0.16), though these differences were not statistically significant.

### Baseline DASH Diet Scores

DASH adherence was poor overall across all participants, with a mean score of 3.8±1.9 out of 9 possible points (42% adherence). DASH scores were similar between groups: 3.7±1.7 in the control group versus 3.9±2.1 in the intervention group (P = 0.74) (**Table 3**). Individual DASH component scores showed consistently low adherence across key nutrients (**Figure 1**). Participants scored below 0.5 out of 1.0 for most components, including calcium (0.3), cholesterol (0.33), fiber (0.46), magnesium (0.46), potassium (0.28), and sodium (0.43). Only total fat (0.51) and saturated fat (0.53) targets approached adequate levels.

**Figure I.**
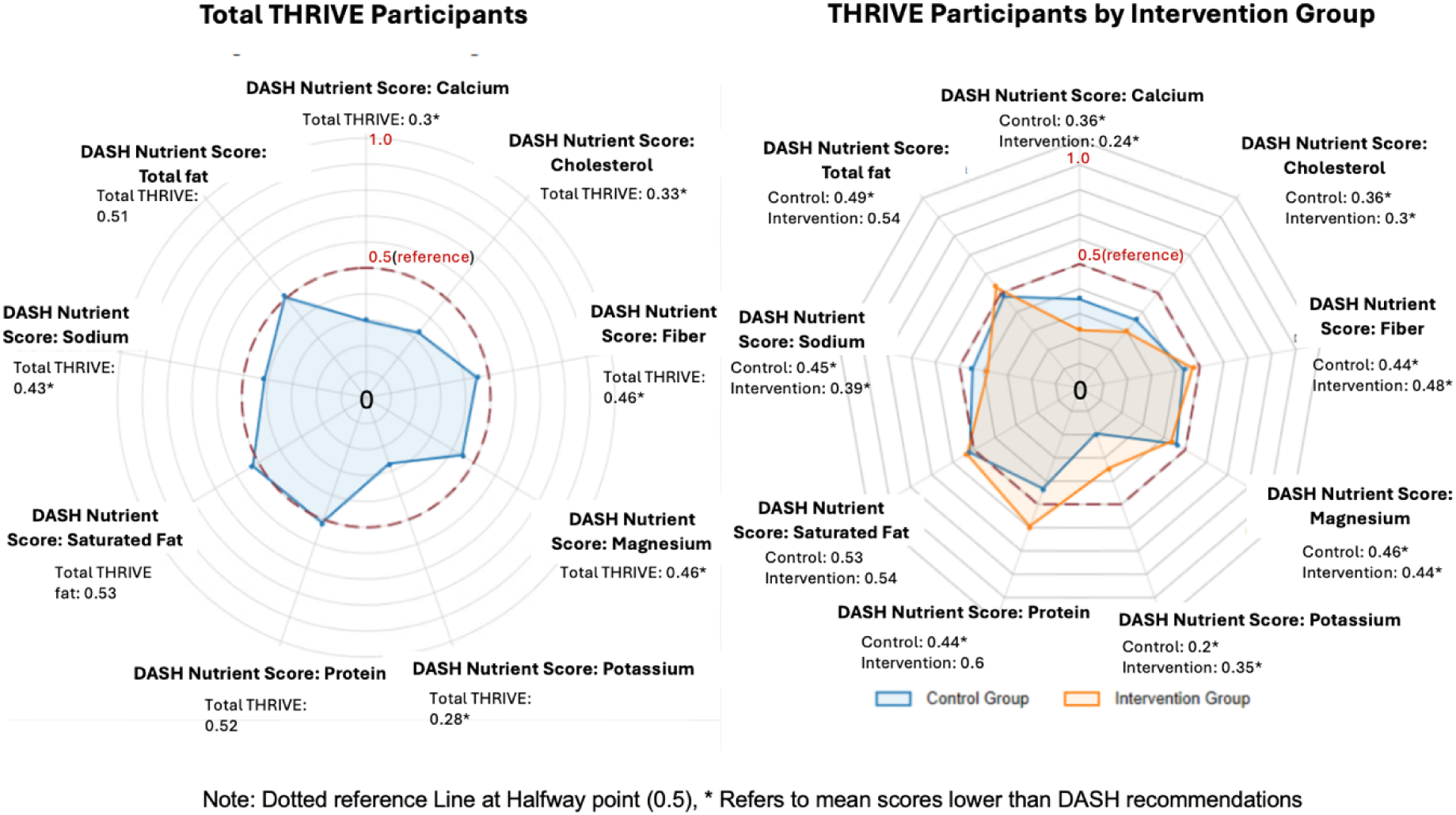
Radar Plot of DASH Scores by Nutrient Targets.

When categorized by adherence level using validated cut-points, 58% of participants had low DASH adherence (scores <4.5) (**Figure 2**). The control group had slightly higher rates of low adherence (62.5% versus 52.5% in the intervention group), but this difference was not statistically significant (χ² = 2.42, P = 0.119).

**Figure II.**
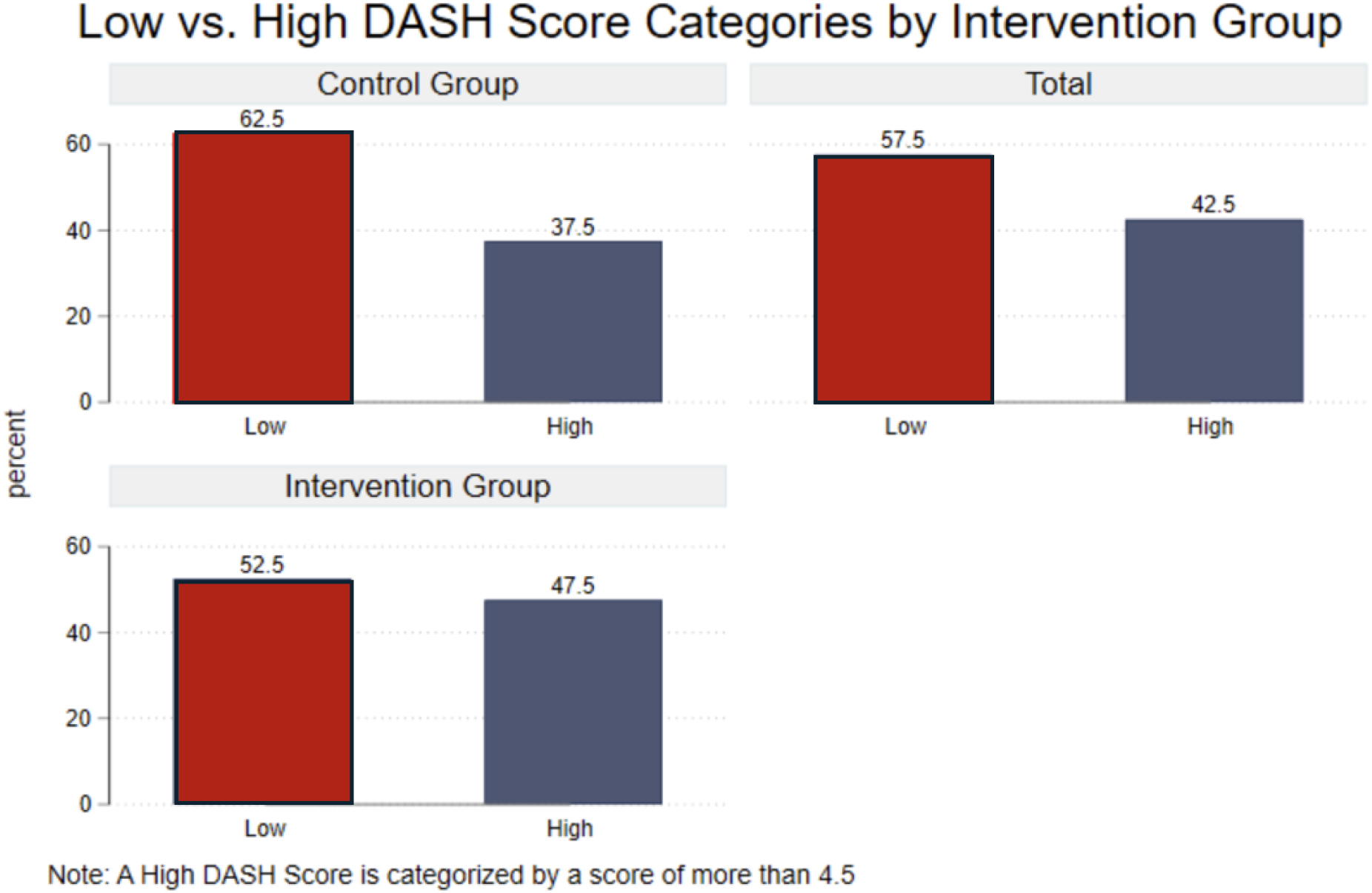
Percentage of Low vs. High DASH Scores.

## DISCUSSION

The THRIVE Food-is-Medicine pilot clinical trial aimed to test the feasibility of combining produce prescriptions, dietitian coaching, and adaptive messaging to improve DASH diet adherence in minoritized populations within HFPAs. Of the 80 participants enrolled, 44.6% of participants had Stage 2 Hypertension, with an average blood pressure of 138.1/82.6 mmHg, reflecting successful recruitment of individuals at high-risk for cardiovascular disease.

These baseline findings underscore the critical need for Food-is-Medicine interventions as defined by the American Heart Association: “the provision of healthy food such as medically tailored meals, medically tailored groceries, and produce prescriptions to treat or manage specific clinical conditions in a way that is integrated with and paid for by the health care sector.”^51^ The THRIVE population exemplifies the target demographic for such interventions, i.e., individuals with clinical conditions (hypertension) who face structural barriers to accessing healthy foods. The intersection of clinical risk and social vulnerabilities observed in our participants aligns with growing recognition that Food-is-Medicine interventions must address both individual health needs and broader social determinants. As noted in recent policy discussions, Food-is-Medicine programs are distinct from, yet complementary to, traditional food assistance programs like SNAP, as they target specific clinical conditions rather than general food insecurity.^19^ Our findings of 36% food insecurity and 37.8% nutrition insecurity alongside 45% Stage 2 hypertension illustrate this dual need; participants require both food access support and medically-tailored nutrition interventions.

The poor DASH adherence scores (mean 3.8/9) despite high educational attainment highlight that knowledge alone is insufficient without addressing structural barriers. This supports the THRIVE approach, which combines produce prescriptions with dietitian coaching and cultural tailoring,^23^ recognizing that effective Food-is-Medicine interventions require culturally responsive strategies that go beyond simple food provision. At baseline, there were numerous cardiometabolic risk factors, including elevated BMI, low DASH nutrient scores, and metabolic irregularities. Most were immigrants, averaging nearly 20 years of residence in the U.S. Half held a master’s degree, yet over 50% earned less than $50,000 annually. This education-income gap is well-documented among immigrant and racially minoritized populations, potentially reflecting structural barriers such as employment discrimination, underemployment, and credential transferability issues.^52,53^ Healthcare access was also limited among participants, with only 77.8% having health insurance coverage, which is lower than the national average of 86%.^54^ This disparity likely contributes to low healthcare utilization. While many participants had received a hypertension diagnosis, only half reported owning a blood pressure monitor at home. This highlights broader inequities in preventive care and chronic disease management among underserved populations.^7^

Dietary recall data collected were consistent with the classification of participants’ neighborhoods as HFPAs. Limited access to affordable, nutrient-rich food is a well-known barrier to diet adherence, particularly in low-income, racially minoritized communities.^55^ High costs, time constraints, limited access to preferred cultural foods, limited access to dietitian coaching, and perceived taste and accessibility issues often prevent individuals from adopting healthier eating patterns, even among those who want to improve their diet.^56^

Low DASH scores across both the control and intervention groups (average 3.83/9) reflect national patterns, particularly among Black and Hispanic adults. Less than 20% of U.S. adults meet optimal DASH adherence, with adherence even lower among low-income, food-insecure, and racially minoritized populations.^38,57,58^ These disparities are likely shaped by structural determinants of health, including food literacy, affordability, availability, and limited culturally appropriate dietary resources.^8,59^ Thus, there have been calls for culturally tailored adaptations to the DASH diet to improve feasibility and acceptance in racially and ethnically diverse communities. Notably, while added sugar and lactose intake differed slightly between groups, all other baseline characteristics were similar. Indication of successful randomization and that the observed intervention effects in later phases can be attributed to the intervention itself. Higher added sugar consumption in the control group may reflect increased acculturation to Western dietary patterns, a trend identified in long-term immigrants.^60^

### Limitations

Several limitations should be acknowledged in interpreting these findings. First, the sample size of 80 participants limits the generalizability of the results. The study focused on specific counties in Maryland, which may limit the applicability of these findings to other regions with different demographic or socioeconomic profiles. Additionally, reliance on self-reported data for sociodemographic and dietary measures introduces potential bias or inaccuracies. As these are baseline findings, the intervention’s impact on participants’ dietary behaviors, hypertension, and cardiovascular health remains to be assessed. Despite these limitations, the baseline data offer valuable insights into the target population’s needs and characteristics, providing a strong foundation for evaluating the effectiveness of the THRIVE intervention in improving DASH diet adherence and reducing hypertension disparities in future phases of the study.

### Perspectives

Overall, the baseline data provides essential context for understanding the THRIVE study population and the factors that influence their diet and health outcomes. These findings underscore the importance of evidence-based, culturally responsive, and accessible cardiovascular interventions for populations in HFPAs with limited healthcare access.

## CONCLUSION

The THRIVE Food-is-Medicine Trial has established a strong foundation for understanding the unique needs and challenges of Black and Hispanic populations with hypertension living in Healthy Food Priority Areas. The baseline data highlight the high prevalence of risk factors for cardiovascular disease, such as high blood pressure, obesity, and food insecurity, as well as the importance of addressing social determinants of health. The overall DASH Diet nutrient scores were low for both groups, suggesting a need for a culturally competent intervention to increase DASH adherence. The study’s success in randomizing participants ensures that future results will reflect the impact of the intervention on these factors. As the intervention progresses, further analysis will determine whether the combination of produce prescriptions, dietary coaching, and adaptive messaging can lead to meaningful improvements in DASH diet adherence and hypertension management in this high-risk population. These findings have important implications for health policy and healthcare delivery. The documented intersection of clinical risk, food insecurity, and structural barriers in this population supports the need for integration of Food-is-Medicine interventions within healthcare systems, particularly for communities residing in areas with limited healthy food access.

Policy makers should consider several key actions based on these findings: expanding Medicaid coverage for Food-is-Medicine interventions in states with high proportions of residents living in food priority areas; investing in community health worker programs that can provide culturally competent nutrition education and care coordination; and developing reimbursement mechanisms that support multi-component interventions rather than isolated clinical services. Additionally, federal nutrition assistance programs should be strengthened and protected as essential complements to clinical Food-is-Medicine interventions.

The evidence for sustained investment in Food-is-Medicine research and implementation is compelling. Continued research should focus on identifying the most cost-effective intervention components, optimal targeting strategies for high-risk populations, and scalable delivery models that can be implemented across diverse healthcare systems. Policymakers must recognize that addressing cardiovascular health disparities requires coordinated investment in both clinical interventions and the social infrastructure that supports healthy communities. Only through sustained, evidence-based Food-is-Medicine interventions can we meaningfully reduce cardiovascular health disparities and enhance population health outcomes across underserved communities nationwide.

## Data Availability

The datasets generated and/or analyzed during the current study are not publicly available but are available from the corresponding author upon reasonable request.

## Acknowledgments

The THRIVE Pilot Study team is incredibly grateful for the participants who agreed to contribute to this study. We are also grateful to the Johns Hopkins ICTR Nutrition Team, the Yuzu Labs Studypages team, the Medi team, the Care for Your Health team, the Mobile Med team, and the Community Farm Share team for their contributions to the implementation of the study. We extend our gratitude as well to the community centers that permitted us to screen and collect data from participants at their locations: Kingdom Fellowship AME Church, Harvest Intercontinental Church, Church of Pentecost Gaithersburg Central, Church of Pentecost PIWC Gaithersburg, Bread of Life Fellowship, First Alliance Church, People’s Baptist Community Church, East County Regional Service Center, and Mongomery County Volunteer Center.

## Disclosures

### Ethics approval and consent to participate

Ethical approval was obtained from the Johns Hopkins University School of Medicine Institutional Review Board (IRB00427492). Additionally, this study was registered in clinicaltrials.gov (NCT06257550)

### Consent for publication

This is not applicable as all data presented here is de-identified.

